# Monitoring of circulating monocyte HLA-DR expression in a large cohort of intensive care patients: relation with outcome and secondary infections

**DOI:** 10.1101/2020.05.01.20087338

**Authors:** C. de Roquetaillade, C. Dupuis, V. Faivre, AC. Lukaszewicz, C. Brumpt, D. Payen

## Abstract

**Background:** post-injury acquired immunodepression (AID) is frequently assessed by the diminished expression of Human Leukocyte Antigen-D Related on circulating monocytes (mHLA-DR). The relation with mortality and the occurrence of ICU-acquired infections (IAI) requires confirmation in large cohorts of patients. This study tested in a large number of ICU patients from a single center the association of mHLA-DR with mortality and secondary infections.

**Methods:** This prospective, observational study in a surgical ICU of a French tertiary hospital reports mHLA-DR measurements (fixed flow cytometry protocol) done 1st within the 3 days post-admission and 2nd after the 7th day. The other collected parameters were: the SAPS II and SOFA scores, sex, age, comorbidity, mortality and ICU-acquired infections. The associations between mHLA-DR and outcomes were tested by adjusted Fine and Gray sub-distribution competing risk models.

**Findings:** 1053 patients were subdivided into 4 subgroups depending on the main motif for admission. Overall, 151 patients (14.3%) died in the ICU with an independent association with the amplitude of the first mHLA-DR decrease (HR = 0.71 [0.57; 0.95], p < 0.01); 592 patients had a 2nd mHLA-DR measurement of whom 223 patients (37.7%) complicated by IAI. These patients had a lower mHLA-DR than other patients (mHLA-DR = 9.0 log vs. 9.3 log, p < 0.01). IAI occurrence was independently associated with first, the 2nd mHLA-DR level regardless the initial severity (HR = 0.66 [0.51; 0.84], p = 0.001) and second, with the slope between the 1st and 2nd values (HR = 0.62 [0.43; 0.89], p = 0.009).

**Interpretation:** the association between the early mHLA-DR expression and ICU mortality does not improve the prediction given by the severity scores. The persistence or a decrease of low mHLA-DR expression are independent and reliable predictors of ICU-acquired infection.

**Funding:** no financial interest

## INTRODUCTION

Acute systemic inflammation characterizes the vast majority of the patients admitted in intensive care (ICU) related to various acute injuries such as sepsis^1^, major surgical procedure^2^, severe polytrauma^3^ and medical diseases^4^. The prediction of the prognosis based on the level of severity scores APACHE 2, SAPS II or SOFA appears difficult to improve by the addition of biomarkers, despite an impressive list of reported biomarkers.^5^ This may results from the heterogeneity of the patients and from the interplay with the host fitness of complex inflammatory pathways, particularly when co-morbidities and chronic treatments are present. The recent evidence of an early and profound acquired immunodepression (AID) in ICU patients has led to a change in the paradigm of acute inflammation.^6^ This temporal shift of both innate and adaptive immunity towards an AID strongly suggests the need for immune monitoring as it is done for other organs in the ICU context. Ideally, this monitoring has to better predict the prognosis and to characterize the risk of ICU-acquired infection (IAI). It may also help the clinician to decide using immunostimulating drugs and to assess their effectiveness avoiding the risk of overtreatment.^7^ Until now, no biomarker has been properly validated to be generalized as an everyday exam. Among the reported biomarkers for the characterization of immune status, the expression of human leucocyte antigen-D related on circulating monocyte (mHLA-DR) has gained credence during the past decade. This functional marker of the Major Histocompatibility Complex II characterizes the functionality of the "immune synapse" between innate and adaptive immunity. Monocytes with low HLA-DR expression are know to be unable to mount an antibacterial response or to properly present antigens to T cells. ^1–4,8–10^ The HLA-DR expression on monocytes is quantified by flow cytometry, which is a robust and reproducible method which makes it suitable for routine examination.^11^ The validation of mHLA-DR expression as a biomarker for mortality prediction is not clearly established lacking reports on large cohorts and/or multicentric studies.^8,10^ The relation between persisting low mHLA-DR expression and the risk of subsequent IAI had been repeatedly shown in limited cohorts, with no clear definition of the minimum threshold for risk definition, nor determination of the most suitable parameter between the intensity of downregulation and its time duration.^8^ In this perspective, our study aimed to: - report the values of mHLA-DR measured early after admission and after 7 days in different life-threatening conditions, investigate the predictive value for prognosis and occurrence of IAI in a single-center large cohort of ICU patients; - compare the profile of AID according to the clusters of motifs for ICU admission.

## MATERIAL AND METHODS

This study was approved by Cochin Hospital Ethics Committee (# CCPPRB 2061, Assistance Publique Hôpitaux de Paris). The mHLA-DR blood tests did not require the patient’s informed consent since it was performed on the remaining routine blood samples with a guarantee to use the data after their anonymization.

### Study design and population

From 2013 to 2015, we prospectively conducted a single-center survey on mHLA-DR monitoring in a large cohort of patients admitted to the surgical ICU. The present cohort was listed from the database when they had at least one measurement of mHLA-DR performed within the first 3 days following ICU admission. Four main clusters of life-threatening conditions were defined according to the motif of admission: 1/ Sepsis, defined by the criteria of the American College of Chest Physicians/ Society of Critical Care Medicine;^12^ 2/ Neurologic disorders, related to acute brain injury such as hemorrhagic or ischemic stroke; isolated severe brain trauma; post-neurosurgery; 3/Post-surgery, related to major abdominal surgery, orthopedic surgery or others; 4/ Miscellaneous etiologies including respiratory failure, hemorrhagic shock from gastrointestinal bleeding or obstetric emergencies. The primary objective was to test whether mHLA-DR measurements at admission or after 5 to 7 days could predict day 28 mortality. A similar investigation was performed to predict early death (before day 7) and late death (from day 7 to day 28). The second goal was to test whether mHLA-DR value was associated with the occurrence of IAI. IAI was diagnosed using the classic definition:^13,14^ a new-onset infection starting at least 48 hours after ICU admission, which motivated a new antimicrobial therapy. The likelihood of infection motivating the clinical decision to administer antibiotics was classified as none, possible, probable and definite.

### Circulating Monocyte HLA-DR measurements (mHLA-DR)

The quantification of the expression of HLA-DR on monocytes was assessed using the number of antibodies per cell (AB/C) by flow cytometry (FACS Canto II instrument, FACS Diva software, Becton Dickinson, San Jose, CA USA) as previously described^8^ (see detailed protocole in the supplementary appendix). In our center, the median and IQ range of mHLA-DR expression in healthy people (n = 13) was a median log mHLA-DR value of 10.6 (IQR: 10.5 - 10.7). The first blood sample and measurement of mHLA-DR was performed within the first 3 days after admission. The 2^nd^ measurement of mHLA-DR was obtained at fixed days (Monday or Thursday) until the patient’s discharge or death. Only patients having two measurements were analyzed to investigate the relation with ICU-acquired infection. We used the threshold of AB/C <8000 to define "low mHLA DR" corresponding to the acquired immune suppression as previously proposed (NCT02361528).^15^

### Statistical analysis

Baseline characteristics (age, sex, comorbidities) were collected on admission. SAPS II and SOFA scores were computed after 24hrs. The data were described as number and percentage for categorical variables and median (interquartile range (IQR)) for continuous variables. Comparisons relied on the Fisher exact test or *χ*^2^ test for categorical data and the Kruskal-Wallis or Wilcoxon test for continuous data. Because of non-linearity, all the mHLA-DR values were log-transformed. Age, SAPS II score and SOFA score were categorized based on the median value. A p-value of less than 0.05 was considered statistically significant.

The association between mHLA-DR measurements and outcomes was assessed using adjusted Fine and Gray subdistribution competing risk models.^16^ For that purpose, first, the subdistribution hazard of mHLA-DR measurements on death at day 28 took into account the competing ICU discharge. The subdistribution hazard of mHLA-DR measurements on the occurrence of IAI at day 28 was made considering the competing ICU death and ICU discharge. For each model, risk factors for the different outcomes were first researched by univariate analyses. The covariates tested into the models were the following: age, motif of admission, SOFA and SAPS II score on admission. Then, the variables yielding P-values <0.2 in univariate analysis were entered into a multivariate model using a backward selection, with p < 0.05 considered significant. The mHLA-DR measurement was forced into all the models. Results were expressed as sub-distribution hazard ratios (sHR) with their 95% confidence intervals (95% CIs). We performed internal validation using a bootstrapping procedure, which was done by taking a large number of samples of the original one. This technique provides nearly unbiased estimates of the confidence intervals (CI) of the odds ratio (OR) of the independent covariates. The missing data were handled via linear interpolation. All analyses were performed using SAS software, version 9.4 (SAS Institute Inc., Cary, North Carolina).

## RESULTS

### Description of the cohort

Among the screened 1766 patients admitted in our ICU during the study period, 1053 patients were finally analyzed (for Flow-chart, see supplementary figure S1). Patients were excluded when: the discharge was before day 4 (n=576); the patients died before the first mHLA-DR measurement (n = 86); a missing 1^st^ mHLA-DR measurement within the first 3 days (n = 51). Motifs for admission were: brain injury (n= 384, 36.5%); sepsis (n = 255, 24.2%, including n=77, 7.3% septic shock); major surgery (n = 80, 7.6%); miscellaneous diagnoses (n = 334, 31.7%) (Table 1). Overall, the mortality rate at 28 days was 14.3% (n= 151) occurring at the median 6^th^ day (IQR [3; 14]). 82 patients (7,9%) died before the 7^th^ day. The mortality rate from day 7 to day 28 was 12% (n=60/499). The median of the 1^st^ mHLA-DR expression was 9.2 log (IQR 8.7 - 9.7). According to the protocol, the delay between admission to first mHLA-DR measurement was 2 [1; 3] days. The number of patients with mHLA-DR < 8000 AB/C (low mHLA-DR) was 38.3% (n = 403). The first mHLA-DR was significantly lower among patients admitted for sepsis and after major surgery compared to other subgroups (supplementary table S1). mHLA-DR expression values for all clusters were lower than those obtained from healthy volunteers (supplementary figure S2).

**Table 1:**
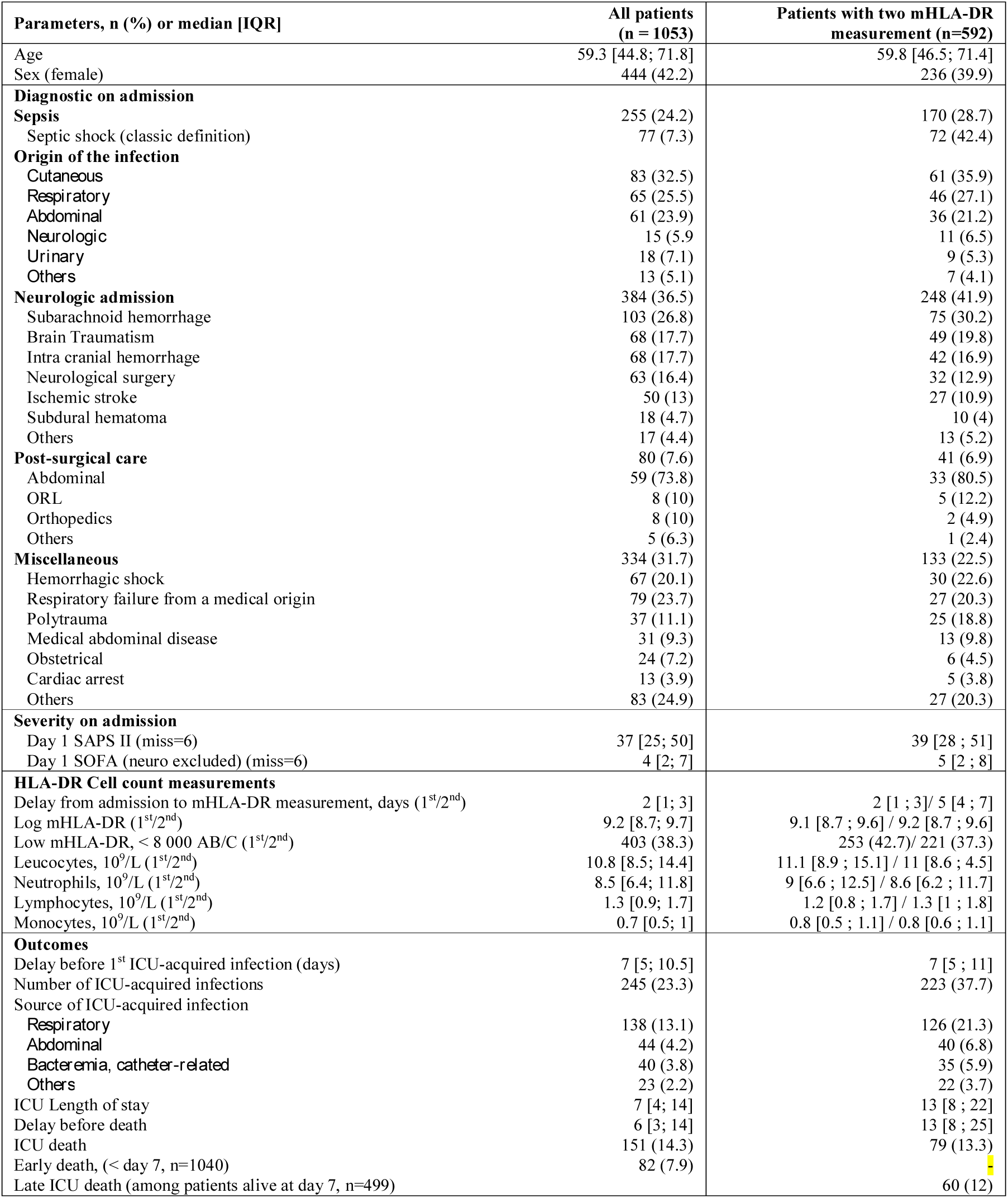
Characteristics of the global cohort and patients having 2 measurements of mHLA-DR IQR: interquartile; ICU: Intensive care Unit-SAPS: Simplified Acute Physiology Score-SOFA: Sequential Organ Failure assessment; NLCR: Neutrophil-to-lymphocyte count ratio; mHLA-DR: monocytic Human Leukocyte Antigen – antigen D Related; AB/C: Antibody per Cell. For diagnostic at admission, data are expressed as percentage within the subgroups

The second mHLA-DR measurements performed within the first week was obtained in 592 patients (Table 1) mainly from patients admitted for brain injury (n=248, 41.9%) and sepsis (n = 170, 28.7%) because they had a longer ICU length of stay (supplementary table S1). The 2^nd^ measurement was not performed in the remaining patients because they died before the 2^nd^ measurement (n=72) or were discharged (n=389) (supplementary figure S1). One IAI was diagnosed in 223 patients (37.7%) (Table 1). The median delay between admission and first ICU-acquired infection episode was 7 days (IQR [5; 11]). The distribution of these infections were predominantly due to the lung (n= 126, 56.5%), the abdomen (peritonitis, biliary tract) (n=40, 17.9%) and bacteriemia/catheter-related infections (n= 35, 13.3%). The rate of these IAI was higher after major surgery (n=28, 48.8%) and brain injury (n=109, 43.95%) than after sepsis (n=54, 31.8%, p<0.01) (supplementary table S2).

### Association between mHLA-DR expression and mortality

In the whole cohort, patients who died were older (p<0.01) with a more severe SAPS II on admission (p<0.01) and a lower mHLA-DR expression than survivors (p<0.01) with a higher proportion of patients having a low mHLA-DR expression (Table 2, p <0.01). After multivariate analysis, the independent risk factors for day-28 mortality were severity scores (SAPS II and SOFA) and brain injury as motif of admission (Table S3). After adjustment, the first measurement of mHLA-DR expression was associated with mortality at day 28 (HR = 0.75 95%CI [0.55-0.94], p=0.01, Figure 1). Among patients with two measurements of mHLA-DR, both the first and the second measurement were associated with ICU mortality (HR 0.63 [0.43; 0.92], p=0.02 and 0.53 [0.4; 0.92], p<0.01 respectively). A low value for the 2^nd^ measurement of mHLA-DR (HR 2.52 [1.56; 4.09], p<0.01) and a decreasing slope between the first and the second measurement (HR 2.13 [1.33-3.45], p<0.01) were independently associated with mortality (Figure 1).The 1^st^ mHLA-DR values did not differ between patients who died early and patients dying after day 7 (p=0.31) (supplementary figure S3). For patients with an under-median SAPS II, the 1^st^ mHLA-DR expression was associated with mortality (late death) but not the 2^nd^ mHLA-DR measurement (supplementary table S4). For patients having a supra-median SAPS2, the 1^st^ mHLA-DR expression did not differ between survivors and dead patients (early death). Only the 2^nd^ mHLA-DR was associated with mortality (late death; p<0.01; supplementary table S5). In the septic group, only the negative slope between the two mHLA-DR measurements was associated with late mortality (p = 0.02). Among patients suffering from "neurological" injury, both the 1^st^ and the 2^nd^ mHLA-DR measurements were associated with early and late mortality (p< 0.01).

**Table 2.** Baseline Characteristics and Outcome of Patients Admitted with Sepsis Stratified according to day-28 mortality and to Development of ICU-Acquired Infection or Not IQR: interquartile; ICU: Intensive care Unit-SAPS: Simplified Acute Physiology Score-SOFA: Sequential Organ Failure assessment; NLCR: Neutrophil-to-lymphocyte count ratio; mHLA-DR: monocytic Human Leukocyte Antigen – antigen D Related. AB/C: Antibody per Cells. For diagnostic at admission, data are expressed as percentage within the subgroups

|  | Alive at Day 28 | Deceased at day 28 | P value | No ICU-acquired infection | ICU-acquired infection | P value |
| --- | --- | --- | --- | --- | --- | --- |
| **Parameters, n (%) or median [IQR]** |  |  |  |  |  |  |
| **Whole Cohort** |  |  |  | **Two measurements cohort** |  |  |
| Number of patients | 902 | 151 |  | 364 | 133 | . |
| Age | 57.7 [43.3 ; 69.9] | 67 [55.9 ; 80.7] | <0.01 | 60.6 [47.9; 72.4] | 57.6 [45.8 ; 71.1] | 0.29 |
| Sex (female) | 375 (41.6) | 69 (45.7) | 0.34 | 154 (42.3) | 47 (35.3) | 0.16 |
| **Diagnostic on admission** |  |  | 0.1 |  |  | 0.13 |
| Sepsis | 210 (23.3) | 45 (29.8) |  | 114 (31.3) | 38 (28.6) |  |
| Neurologic admission | 326 (36.1) | 58 (38.4) |  | 138 (37.9) | 60 (45.1) | . |
| Post-surgical care | 74 (8.2) | 6 (4) |  | 21 (5.8) | 12 (9) | . |
| Miscellaneous | 292 (32.4) | 42 (27.8) |  | 91 (25) | 23 (17.3) | . |
| **Severity on admission** |  |  |  |  |  |  |
| Day 1 SAPS II | 35 [24 ; 47] | 54 [42 ; 67] | <0.01 | 37 [26 ; 49] | 40 [32 ; 51] | 0.01 |
| Day 1 SOFA (neuro excluded) | 4 [1 ; 6] | 8 [6 ; 12] | <0.01 | 4 [2 ; 8] | 6 [3 ; 9] | <.01 |
| **mHLA-DR, Cell count measurements** |  |  |  |  |  |  |
| Time from admission to mHLA-DR measurement, days | 2 [1 ; 3] | 1 [1 ; 3] | 0.04 | 1 [1 ; 3] | 2 [1 ; 3] | 0.66 |
| Log mHLA-DR | 9.2 [8.8 ; 9.7] | 8.9 [8.3 ; 9.4] | <0.01 | 9.1 [8.7 ; 9.6] | 9.1 [8.7 ; 9.5] | 0.70 |
| Low mHLA-DR, <8 000 AB/C | 318 (35.3) | 85 (56.3) | <0.01 | 155 (42.6) | 56 (42.1) | 0.92 |
| Leucocytes, 10<sup>9</sup>/L | 10.6 [8.4 ; 14.1] | 12.7 [9.1 ; 17.5] | <0.01 | 9.3 [8.8 ; 9.7] | 9.1 [8.6 ; 9.4] | . |
| Neutrophils, 10<sup>9</sup>/L | 8.4 [6.2 ; 11.3] | 10.7 [7.2 ; 14.5] | <0.01 | 9.3 [8.8 ; 9.7] | 9.1 [8.6 ; 9.4] | . |
| Lymphocytes, 10<sup>9</sup>/L | 1.3 [0.9 ; 1.8] | 1.1 [0.7 ; 1.6] | <0.01 | 1.2 [0.8 ; 1.7] | 1.1 [0.7 ; 1.6] | 0.04 |
| Monocytes, 10<sup>9</sup>/L | 0.7 [0.5 ; 1] | 0.8 [0.5 ; 1.1] | 0.44 | 0.7 [0.5 ; 1] | 0.8 [0.5 ; 1.1] | 0.57 |
| **Outcomes** |  |  |  |  |  |  |
| Delay before 1<sup>st</sup> ICU-acquired infection, days | 7 [5 ; 11] | 5 [4 ; 9.5] | 0.13 | - | 9 [6 ; 16] |  |
| Number of ICU-acquired infections | 197 (21.8) | 48 (31.8) | <0.01 | 0(0) | 133(100) |  |
| ICU Length of stay | 7 [4 ; 15] | 6 [3 ; 14] | 0.44 | 9 [6 ; 14] | 24 [17 ; 33] | <.01 |
| Death at day 28 | - | - |  | 39 (10.7) | 23 (17.3) | 0.05 |
| **2 mHLA-DR measurements** |  |  |  |  |  |  |
| Number of patients | 513 | 79 |  |  |  |  |
| Delay before measurement n°1, days | 2 [1 ; 3] | 1 [1 ; 3] | 0.33 | 1 [1 ; 3] | 2 [1 ; 3] | 0.66 |
| Delay before measurement n°2, days | 5 [4 ; 7] | 5 [4 ; 6] | 0.05 | 5 [4 ; 6] | 5 [4 ; 6] | 0.94 |
| Log mHLA-DR (1<sup>st</sup>) | 9.2 [8.7 ; 9.6] | 8.9 [8.1 ; 9.3] | <0.01 | 9.1 [8.7 ; 9.6] | 9.1 [8.7 ; 9.5] | 0.70 |
| Low mHLA-DR 1 | 208 (40.5) | 45 (57) | <0.01 | 155 (42.6) | 56 (42.1) | 0.92 |
| Log mHLA-DR (2<sup>nd</sup>) | 9.3 [8.8 ; 9.7] | 8.6 [8.1 ; 9.3] | <0.01 | 9.3 [8.8 ; 9.7] | 9.1 [8.6 ; 9.4] | <.01 |
| Low mHLA-DR 2 | 169 (32.9) | 52 (65.8) | <0.01 | 116 (31.9) | 58 (43.6) | 0.02 |
| Decreasing slope (mHLA-DR<sub>2</sub><mHLA-DR<sub>1</sub>) | 235 (45.8) | 51 (64.6) | <0.01 | 161 (44.2) | 80 (60.2) | <.01 |
| Slope mHLA-DR | 1.1 [-5.9 ; 8.3] | -3.4 [-12 ; 3.9] | <0.01 | 2 [-5.1 ; 10.1] | -2.7 [-10.8 ; 4.6] | <.01 |
| Low mHLA-DR 1 and Decreasing Slope | 90 (17.5) | 35 (44.3) | <0.01 | 36 (9.9) | 22 (16.5) | 0.04 |

**Figure 1:**
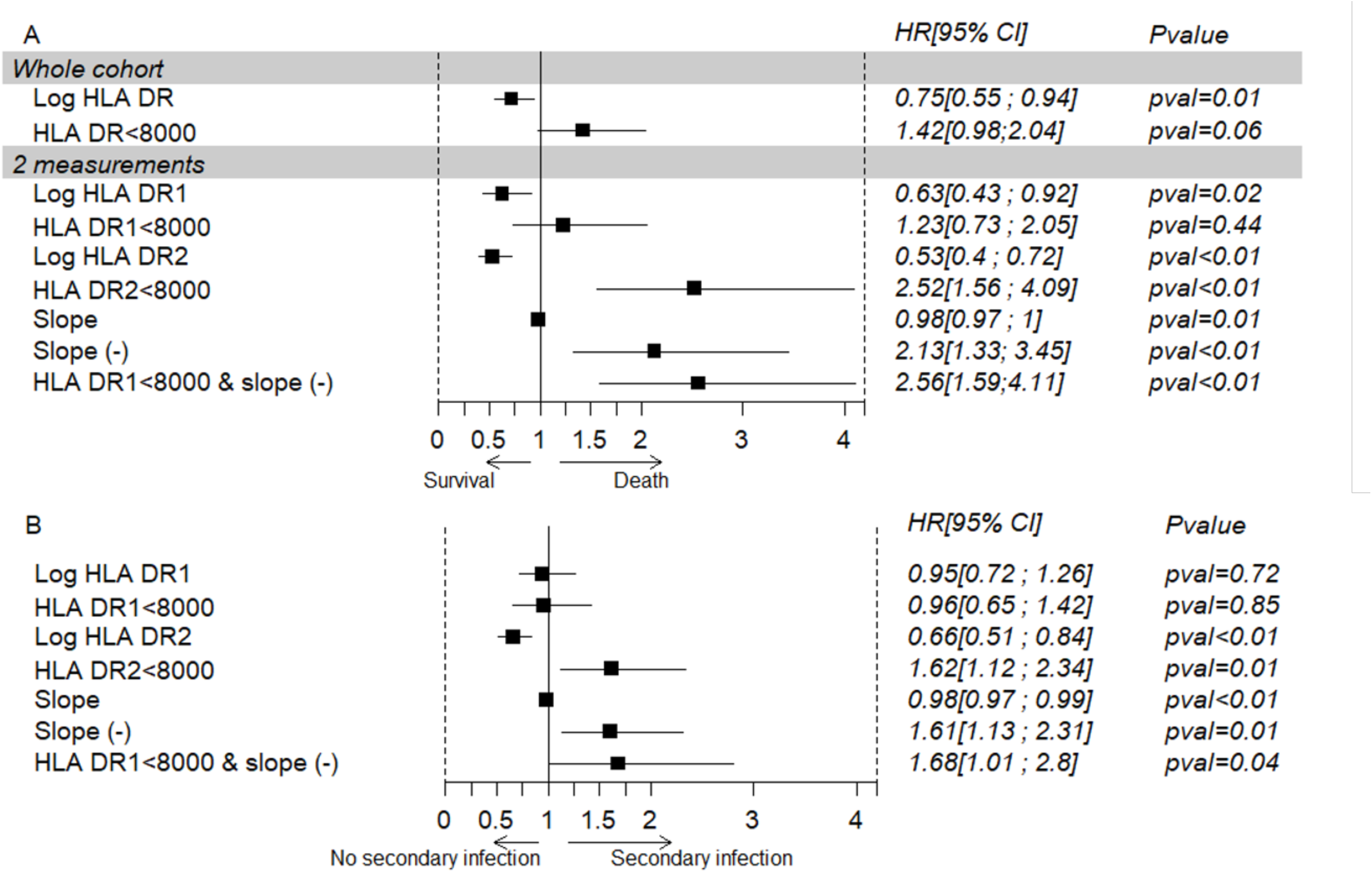
Forrest plot representing the Association between the variables of HLA DR expression (1st measurement, 2nd, and trend of mHLA-DR) and death (A.) or ICU-acquired infections (B.). Multivariate sub-distribution hazard models. HR: Hazard Ratio, CI: Confidence interval, mHLA-DR: monocytic Human Leukocyte Antigen – antigen D Related. For diagnostic at admission, data are expressed as a percentage within the subgroups

### Association between mHLA-DR expression and IAI

Table 2 summarizes the results. The patients having IAI were more severe (SAPS II and SOFA score; p = or < 0.01) with a similar 1^st^ mHLA-DR value (p = 0.70). The 2^nd^ mHLA-DR level (p <0.01), the incidence of a low value 2^nd^ mHLA-DR (p = 0.02) and of negative slope (p <0.01) were significantly different in IAI group compared to the no IAI group. These parameters were independently associated with the occurrence of IAI (Figure 1B and Figure 2). The bootstrap analysis we used confirmed the association between negative slope and occurrence of IAI, with no additional risk when supra-median SAPS II was added in the model (supplementary table S5). The patients having a supra-median SAPS II with a negative slope for mHLA-DR expression had the highest risk for IAI (figure 3). The risk remains higher for infra-median SAPS II. In the "sepsis" subgroup, the multivariate analysis showed a strong association between negative slope (p <0.01) or low 2^nd^ mHLA-DR (p <0.01) and occurrence of subsequent IAI (supplementary table S6).

**Figure 2:**
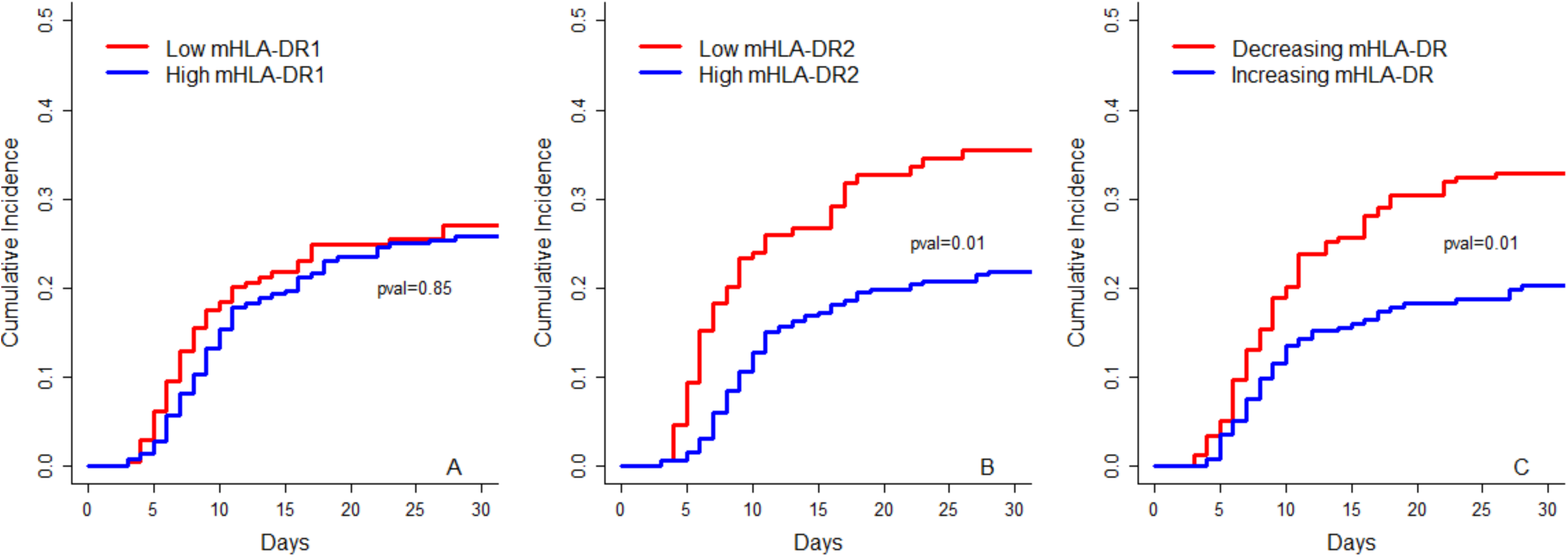
cumulative incidence of the occurrence of nosocomial infection depending on different levels of early mHLA-DR expression (A), second mHLA-DR expression (B) and depending on the trend of mHLA-DR (C) in patients with two measurements (n=592). Low mHLA-DR expression is defined by a level<8000. P-value estimated by a multivariate subdistribution survival model.

**Figure 3:**
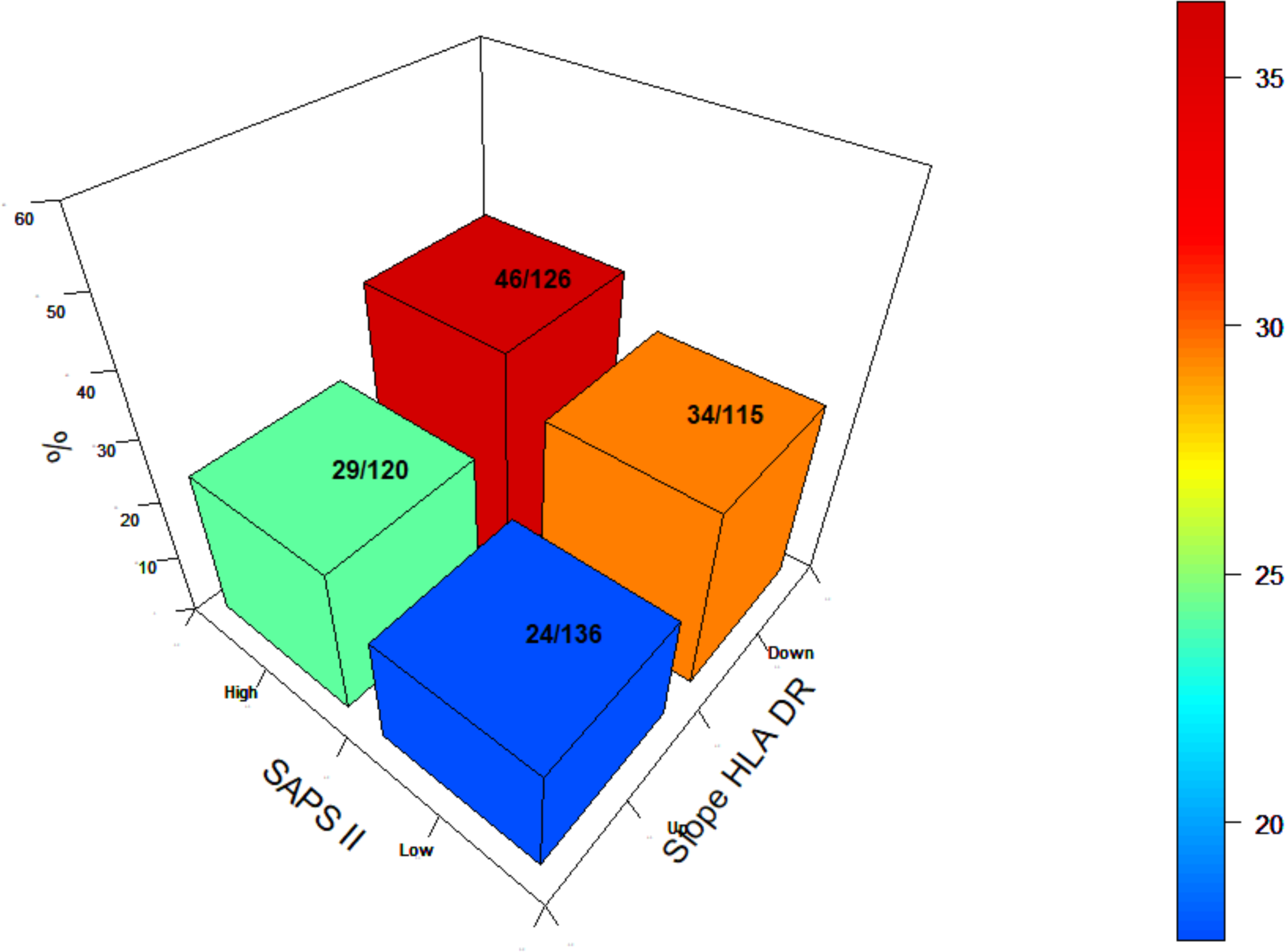
Tridimensional representation of the relation between SAPS II – the slope of mHLA-DR and the occurrence of secondary infection at day 28. Whatever initial gravity, the existence of a decreasing slope between the first and second measurement of mHLA-DR is a strong risk factor for the later occurrence of ICU-acquired infection. SAPS II: Simplified Acute Physiology Score 2; mHLA-DR: Monocytic Human Leukocyte Antigen – Antigen D Related

## DISCUSSION

### Key results

In this monocentric observational large cohort of ICU patients, the monitoring of mHLA-DR during the first days post-admission showed an association between the reduction in mHLA-DR expression and day-28 mortality, but with no additional impact compared to the SAPS II prediction. The persisting low expression of mHLA-DR is the unique significant and independent parameter associated with the development of IAI, which was confirmed by the "bootstrap" testing. The proposed threshold of 8000 events per cell ^15^ is confirmed by the median value found in our study. These results support the interest of the repeated monitoring of mHLA-DR expression during the first days post-admission to identify the patients at risk for IAI.

### Limitations

First, the monocentric evaluation precludes the generalization of the results before the multicentric validation. However, the monocentric design may have advantages: the protocol for caring patients was homogeneous; the method for measuring mHLA-DR expression using the same flow cytometer was fixed and rigorous; the collected clinical contexts covered a large panel of ICU etiologies for systemic inflammation. The results obtained with our large cohort fits well with those previously reported in smaller studies, which confirms the concept of an early post-injury AID for a vast majority of ICU-patients.^8–10^ Second, the flow cytometry method used for this cohort applied the standardized protocol previously proposed by a European task force, especially the normalization for the number of events per cell.^17^ The persistent manual steps for mHLA-DR expression might, however, introduce some signal/noise ratio variations, which could be solved with more automated devices in the future.

### Interpretations

The validation steps for immune biomarkers to predict the risk of complications such as secondary infections require large cohorts and validation by randomized clinical trials. We and others have previously reported similar results in reasonable cohorts of ICU patients.^4, 8, 10, 18^ The present study confirms previously published results obtained among 387 patients admitted in ICU for various primary diagnosis, including sepsis and neurologic patients.^8^ The larger size of the cohort, the fixed protocol for mHLA-DR measurement at fixed time after admission give credence to obtained results. Altogether the previous and the present results confirm the early downregulation of mHLA-DR in different ICU contexts, with a more pronounced acquired immunodepression in septic patients.^6^ The medical community has then reconsidered the paradigm of systemic inflammation, especially for sepsis. Even the underlying mechanisms remain debated, the AID concerns all type of immune cells as neutrophils^19^, monocytes^20^ and lymphocytes^21–23^ and consequently innate and adaptive immunity. Several reviews have conceptualized AID with the potential to propose drugs to "boost" the immune system.^24–26^ Different immune markers to diagnose AID have been proposed to characterize patients’ immune status.^25^ Among these, measurement of monocyte expression of HLA-DR appeared the most pragmatic technique to fulfill the clinical needs: a compatible time for measurement for clinical decision making; a good reproducibility; a numerical value to be compared; a reasonable cost. Before generalization, the relation between the decreased mHLA-DR and outcome had to be clarified. In the present study, the relation between mHLA-DR expression and mortality was analyzed in 1053 patients admitted for different clinical contexts, grouped in 4 clusters. As previously reported, mHLA-DR measured early after admission was lower than control values in healthy subjects for the whole cohort and the 4 clusters of patients. The lowest values were observed both in "sepsis" and "post-operative" patients. If mHLA-DR expression decrease is associated with day 28 mortality, it does not improve the prediction observed with SAPS II. Few data have shown relationship between the amplitude of mHLA-DR reduction and mortality.^8,10^ In contrast with to previous studies, our analyses were provided after adjustments for confounding factors. However, as for previous publications, our conclusions might be limited by the fact that patients dying before day 4 were not enrolled.

The expression of mHLA-DR seems to be a reasonable approach as tested in the present study. The presence of AID occurs soon after admission in almost all types of patients, with a nadir of depression more profound in septic and post-surgical patients. If the reduction of mHLA-DR expression is present, the depth of downregulation does not correlate with the prognosis. Similar patients suffering from similar profound mHLA-DR expression can survive with one etiology and die with others. The global evaluation of mHLA-DR expression in the whole cohort showed a more pronounced median in septic and post-surgical patients, but with a poor prediction of outcome. Some studies have suggested a relation between the depth of mHLA-DR expression and mortality, but with no enrichment of the score predicability.^4,27^ The early 1^st^ measurements remains however important as a reference point for the trending characterization. A later, 2^nd^ measurement, among patients staying in ICU after several days allowed to test the meaning of its value, and the signification of the trend between the 1^st^ and the 2^nd^ values, and to investigate the impact of the duration of mHLA-DR downregulation on outcomes.

Finally, the most promising perspective with monitoring of immune status is its ability to help physician to early identify patients that might benefit from immune restoration intervention. In further randomized controlled trials, screening for patients seems essential and guided by immune monitoring. One more time, our analyses confirms that further study should not focus on ICU-mortality, our observations like others seem to demonstrate a relatively poor correlation with immune status.^28^ In this perspective, we encourage future study to focus on ICU length of stay and/or IAI occurrence more than mortality as a primary endpoint.

## CONCLUSION

the association between the early mHLA-DR expression and ICU mortality does not improve the prediction given by the severity scores. A persisting low or a decrease of mHLA-DR expression in the first 7 days of ICU admission is independently and reliably associated with subsequent IAI.

## Data Availability

Data, analytic methods, and study materials are available to other researchers upon motivated request by email

